# Near Term Predictions of Covid-19 Cases in West Bengal, Maharashtra, Delhi and Tamil Nadu in India Based on Basu Model

**DOI:** 10.1101/2020.07.05.20146910

**Authors:** Santanu Basu

## Abstract

Four states in India account for nearly two thirds of the Covid-19 cases and the numbers are growing fast. A mathematical model which has been recently developed and which has been applied to other countries and regions in the world has been used in this paper to anchor the available data to date and to make near term projections. The projections can be compared to actual data to take steps in the near term in controlling the spread of this disease.

## INTRODUCTION

Since the first confirmation of a Covid-19 patient on Jan 30^th^ in India, the number of confirmed cases has grown to 648,000 and the number of deaths has grown to 19,000 by Jul 4^th^. All dates in this paper are in the year 2020. Almost two thirds of the cases are in four states: West Bengal, Maharashtra, Delhi and Tamil Nadu which are major employment hubs and are densely populated. The new coronavirus, SARS-CoV-2 spreads from person to person [1]. In the most common form of transmission, the virus from an infected person’s nose or mouth enters a healthy person’s nose or mouth if the healthy person is within 6 ft of the infected person without personal protection. The two most effective tools against spreading of the disease are social distancing and use of personal protective equipment, such as a face mask. For convenience we use the term social distancing which is the result of measures such as lockdown, shelter in place, work from home and quarantine. As compared to other large countries, the Indian authorities took prompt and decisive measures in combating the outbreak. A nationwide lockdown was announced on Mar 24^th^ when the total number of Covid-19 cases and deaths were only 519 and 10 respectively. The author believes that this early proactive step saved many lives and resulted in much fewer cases in the first three months of the outbreak than if the lockdown were delayed by even a few weeks.

Since the virus stays in an infected person’s body for about 3 weeks, strict adherence to social distancing and face mask rules purges the virus from the population in low multiples of 3 weeks of residence time. A proof of this method to work is New Zealand in which the Covid-19 disease lasted for only 41 days (Mar 16^th^ to Apr 26^th^, days with cases >5% of peak) [2]. The author has developed a Covid-19 case growth model [3-7], which for brevity will be termed Basu model in this paper. Basu model has been applied to New Zealand and many other countries and states, and it has convincingly shown the effectiveness of reduced transmission rate (through social distancing) in controlling the pandemic.

However a large country such as India faces four primary challenges in implementing the New Zealand method. These are:

- The outbreak starts at different times in different states
- There is population movement across state boundaries
- Different states may have different strategies
- Economic necessity does not allow long durations of social distancing.

It is a point of utmost interest to determine the best course of action given the four challenges as outlined above. After the first lockdown, any successive lockdown needs to be strictly and uniformly enforced nationwide for at least six weeks to simultaneously check the pandemic and to pay the minimum economic price. In this paper, we apply the Basu model to Covid-19 case growth in four states at four different parts of the country to understand the underlying mechanism of case growth and resurgence as social distancing orders are relaxed and tightened. A good quantitative model may help the authorities in charge of controlling the pandemic and restoring normal life.

## OVERVIEW OF THE COVID-19 MATHEMATICAL MODEL

The author developed and documented a mathematical model to predict number of Covid-19 cases and hospital patient loads [3-7]. The reader is encouraged to review the details of the mathematical model in references [3,4,6] which will not be repeated here. The model assumes that starting from a small number of infected people which act a seed, the infection grows by transmission from person to person in proximity. The model keeps track of the number of infected people at various stages of development of the disease through the course of the virus outbreak. The model in its current form is applicable to an isolated region, and work is in progress to include population movement across the region boundaries. Therefore the model predictions for the four states in India studied in the paper are with approximations since in reality there is considerable population movement across state boundaries in these four states. The effect of population movement will soon be included in the model. The model is parametric and transparent so that predictions are based on parameters which are in principle measurable and the predictions can be continuously corrected as more accurate data on parameters become available. Basu model provides greater physical insight into the problem than machine learning and curve fitting type models. The present model has a number of user-supplied parameters, one of which influences the results greatly. It is the transmission factor, which by itself is time dependent and parametrized.

## RESULTS AND DISCUSSION

### West Bengal

Covid-19 case data for West Bengal, India was obtained from [8]. The first case of Covid-19 in West Bengal was confirmed on Mar 17^th^ and the first death was registered on Mar 23^th^. The authorities gave orders to close schools from Mar 14^th^, partial work from home order started on Mar 21^st^ and the nationwide lockdown by the central government was implemented on Mar 24^th^.

Relaxation of the lockdown started on Jun 1^st^ with the allowance of some religious activities and tea shops being open. The offices started opening on Jun 8^th^. During this time, migrant workers were visiting the state by train.

West Bengal data could be fitted well with Mar 24^th^ to be the first date (n_1_) of transmission rate decrease, 15 days for exponential time constant for transmission rate decrease (G) and 7% for the final transmission rate (T_f_). The final transmission rate was increased to 8% starting on Jun 1^st^ (first relaxation order) and to 9% starting on Jun 8^th^ (further relaxation orders). The comparison between the model predictions and actual data on number of cases and deaths are shown in figure 1. The figures demonstrate that the model fits the actual data very well.

**Figure 1.**
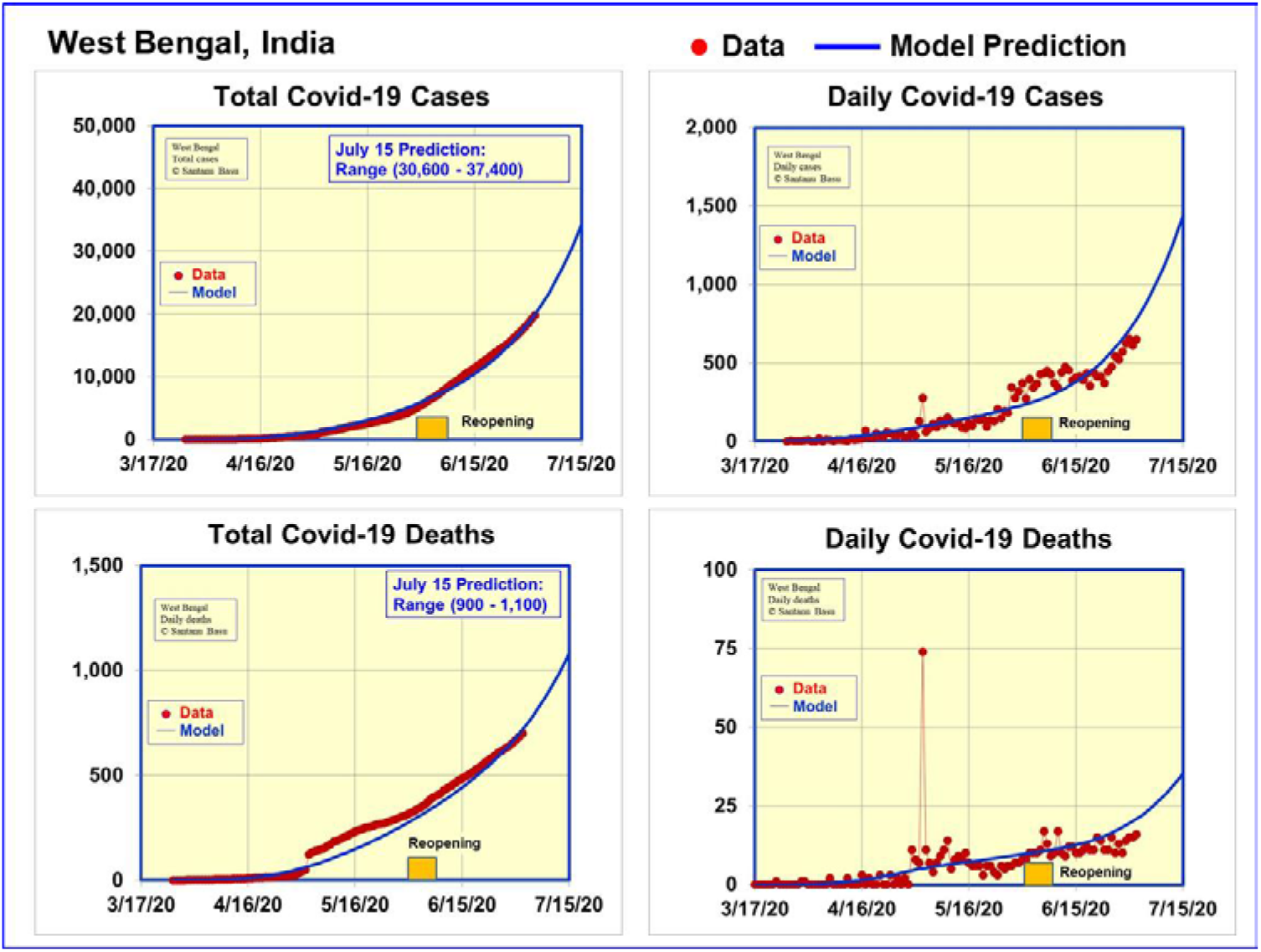
Data until Jul 2^nd^ and model predictions for Covid-19 cases and deaths in West Bengal, India

Figure 1 clearly shows that the positive effect of social distancing and shelter in place has not yet been realized by Jun 1^st^ when the social distancing orders were relaxed. As a result, an upward trend in the number of cases is seen after Jun 1^st^. The model indicates that such level of social distancing is not sustainable as the number of cases will accelerate an upward trend followed by the number of deaths doing the same. It will be advisable to examine the number of cases and deaths on Jul 15^th^ to review the pandemic controlling strategies. The model predicts the number of cases to rise to between 30,600 and 37,400 and the deaths to rise to 900-1,100 by Jul 15^th^.

### Maharashtra

Covid-19 case data for Maharashtra was obtained from [9]. The state had the first case of Covid-19 on Mar 9^th^ and the first death was reported on Mar 17^th^. The authorities gave orders to close places of public gatherings from Mar 14^th^ followed by the nationwide lockdown on Mar 24^th^. Reopening of the state started on Jun 28^th^.

Maharashtra data could be fitted well with Mar 18^th^ to be the first date (n_1_) of transmission rate decrease, 15 days for exponential time constant for transmission rate decrease (G) and 6.6% for the final transmission rate (T_f_). The final transmission rate was increased to 7.6% starting on Jun 28^th^ (first relaxation order). The model predictions are compared with the actual data on number of cases and deaths in figure 2. The figures demonstrate that the model fits the actual data very well.

**Figure 2.**
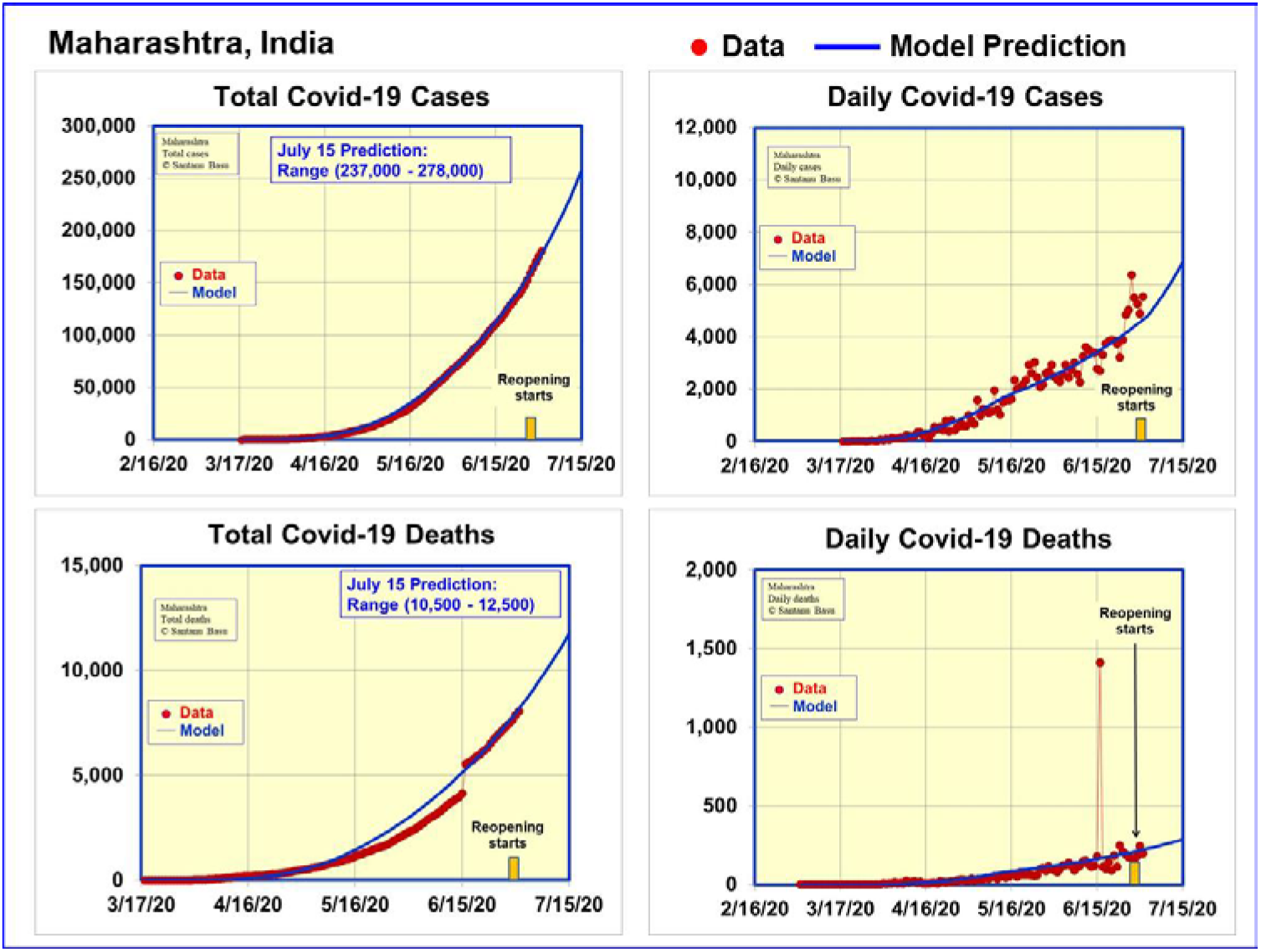
Data and model predictions for Covid-19 cases and deaths in Maharashtra, India

Figure 2 shows that the there is an upward trend in the daily number of cases in late June due to relaxation of social distancing. The model predicts the number of cases to be in the range 237,000 - 278,000 and the deaths to be in the range 10,500-12,500 on Jul 15^th^.

### Delhi

The data for Delhi was obtained from [10]. The first Covid-19 case and death were reported on Mar 2^nd^ and Mar 14^th^ respectively in Delhi. The local authorities gave orders to close public places from Mar 23^rd^ followed by the nationwide lockdown on Mar 24^th^. Gradual reopening started on Jun 10^th^.

Delhi data could be fitted well with Mar 23^rd^ to be the first date (n_1_) of transmission rate decrease, 7 days for exponential time constant for transmission rate decrease (G) and 7.8% for the final transmission rate (T_f_). The final transmission rate was increased to 8.8% starting on Jun 10^th^ with a 15 day time constant. Figure 3 compares the actual data for Delhi until Jun 25^th^ and model predictions until Jul 15^th^, the fit between model and the data is very good.

**Figure 3.**
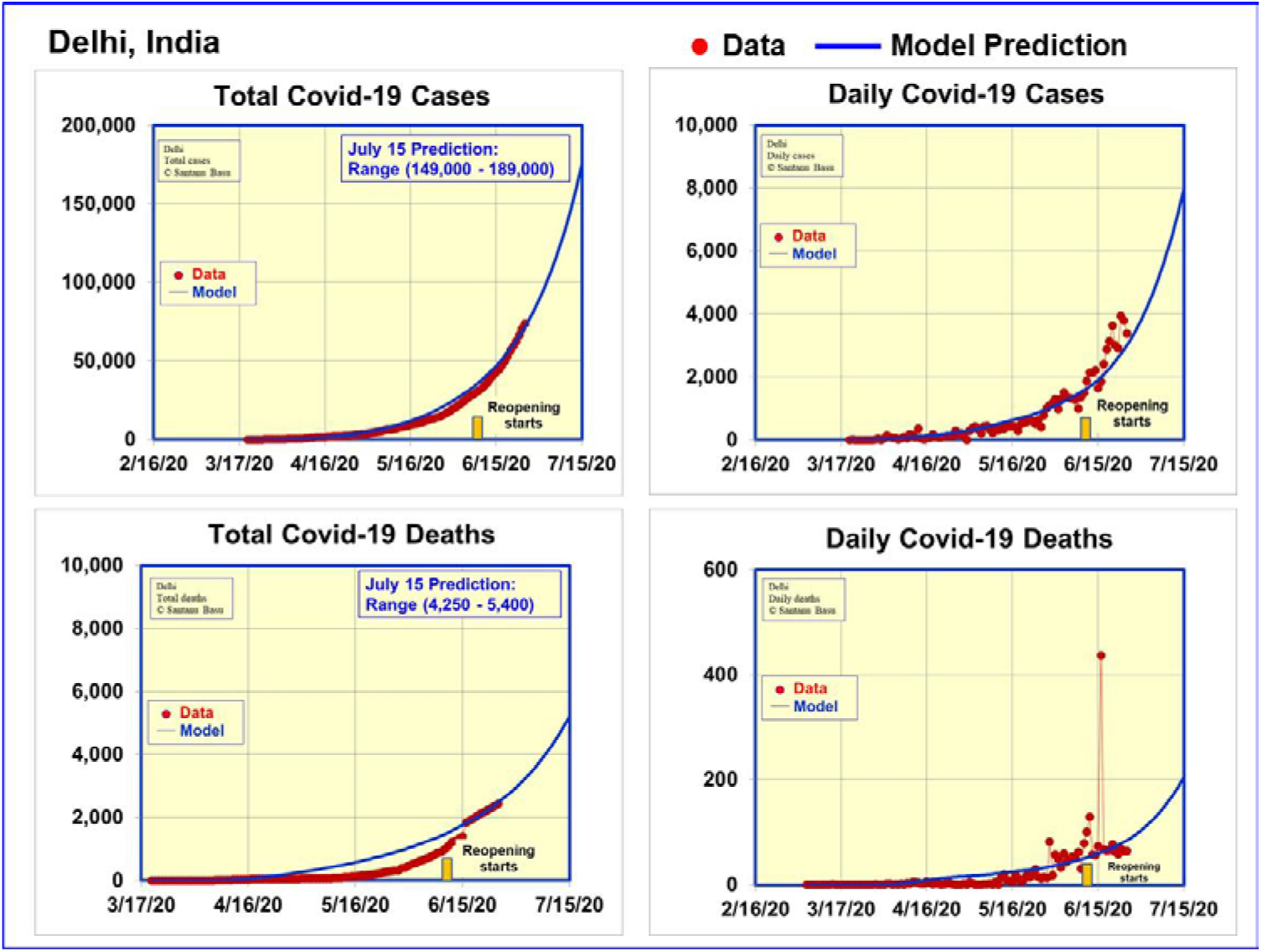
Data and model predictions for Covid-19 cases and deaths in Delhi, India

Figure 3 shows that the number of daily cases was still rising when the social distancing orders were relaxed on Jun 10^th^. As a result, an upward trend in the number of cases is seen after Jun 10^th^. The model predicts the number of cases to rise to be in the range 149,000 – 189,000 and the number of deaths to be in the range 4,250 – 5,400 on Jul 15^th^. The actual data on Jul 15^th^ as compared with the model prediction should allow the authorities to make corrective actions if necessary.

### Tamil Nadu

Covid-19 case data for Tamil Nadu, India was obtained from [11]. The first case of Covid-19 was confirmed on Mar 7^th^ and the first death was reported on Mar 25^th^. Public gathering places started closing on Mar 15^th^ and full lockdown ensued on Mar 22^nd^.

Lockdown relaxations started 50 days later on May 11^th^ followed by further relaxations on Jun 1^st^ and Jun 8^th^. Reopening orders were accompanied by strict guidance on social distancing, face mask use, times of opening, percentage occupancy and cleaning requirements in all states studied in this paper. In some hard hit areas, social distancing orders were maintained.

Tamil Nadu data could be fitted well with Mar 15^th^ to be the first date (n_1_) of transmission rate decrease, 21 days for exponential time constant for transmission rate decrease (G) and 8% for the final transmission rate (T_f_). The final transmission rate was increased to 9% starting on May 22^nd^ with a time constant of 15. The comparison between the model predictions and actual data on number of cases and deaths are shown in figure 4. The figures demonstrate that the model fits the actual data very well.

**Figure 4.**
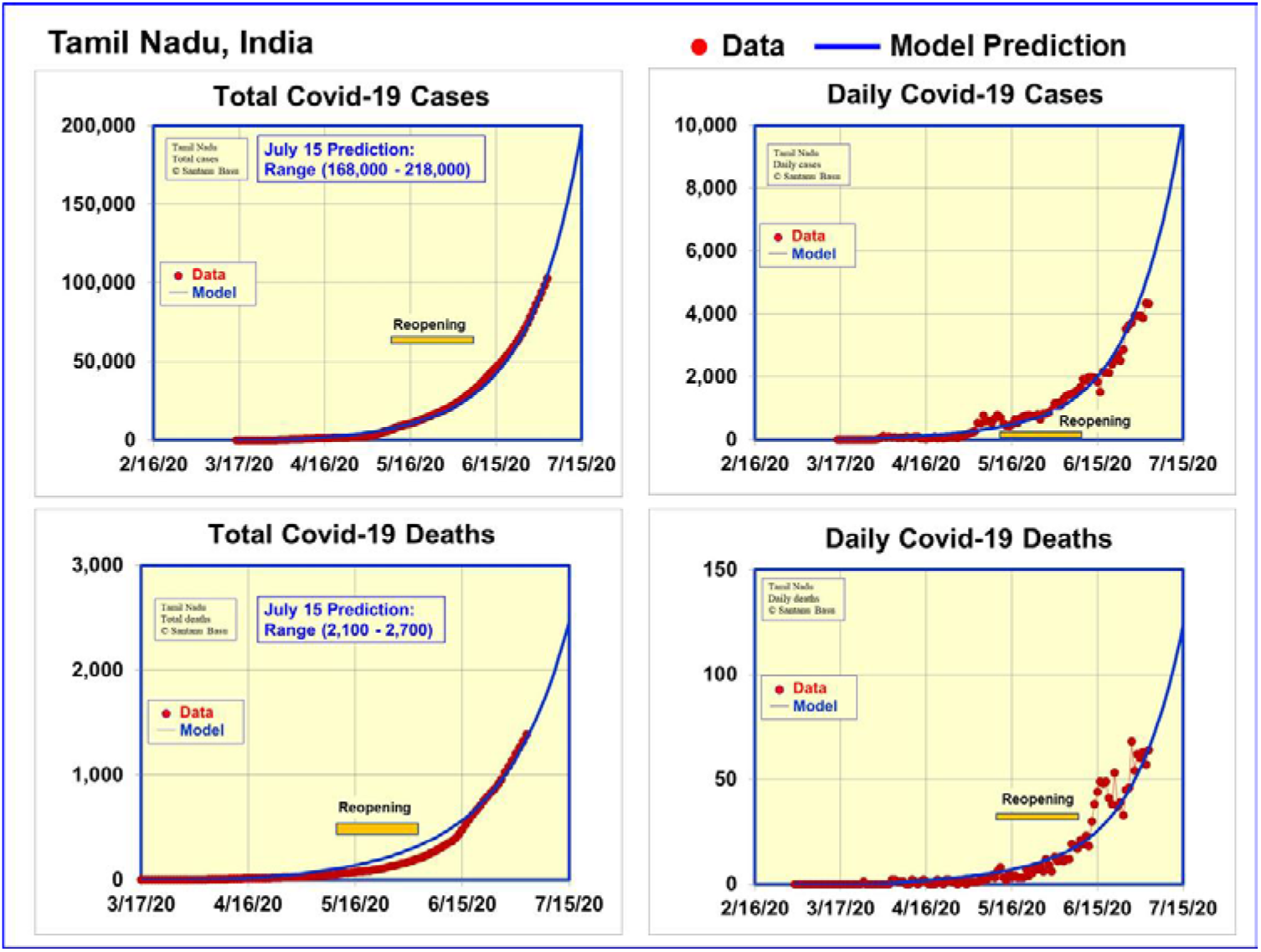
Data and model predictions for Covid-19 cases and deaths in Tamil Nadu, India

Figure 4 shows an upward trend in the number of cases after Jun 1^st^. The model indicates that such level of social distancing is not sustainable as the number of cases will significantly increase in the near term. The model predicts the number of cases to rise to be in the range 168,000 - 218,000 and the deaths to rise to 2,100 - 2,700 by Jul 15^th^.

Figure 5 summarizes the Covid-19 case and death projections for the four states studied in this paper. The predictions are made only up to Jul 15^th^ since the growth of the numbers is dictated strongly by two opposing forces: social distancing orders make the number growth to be slowed, economic activity which is a dire necessity makes the number growth to be accelerated. The balance between these two is being established in these early days after reopening. One interesting feature in the data which we discovered is that the ratio of deaths to cases varies widely from state to state with Maharashtra having 250% more death per case than Tamil Nadu.

**Figure 5.**
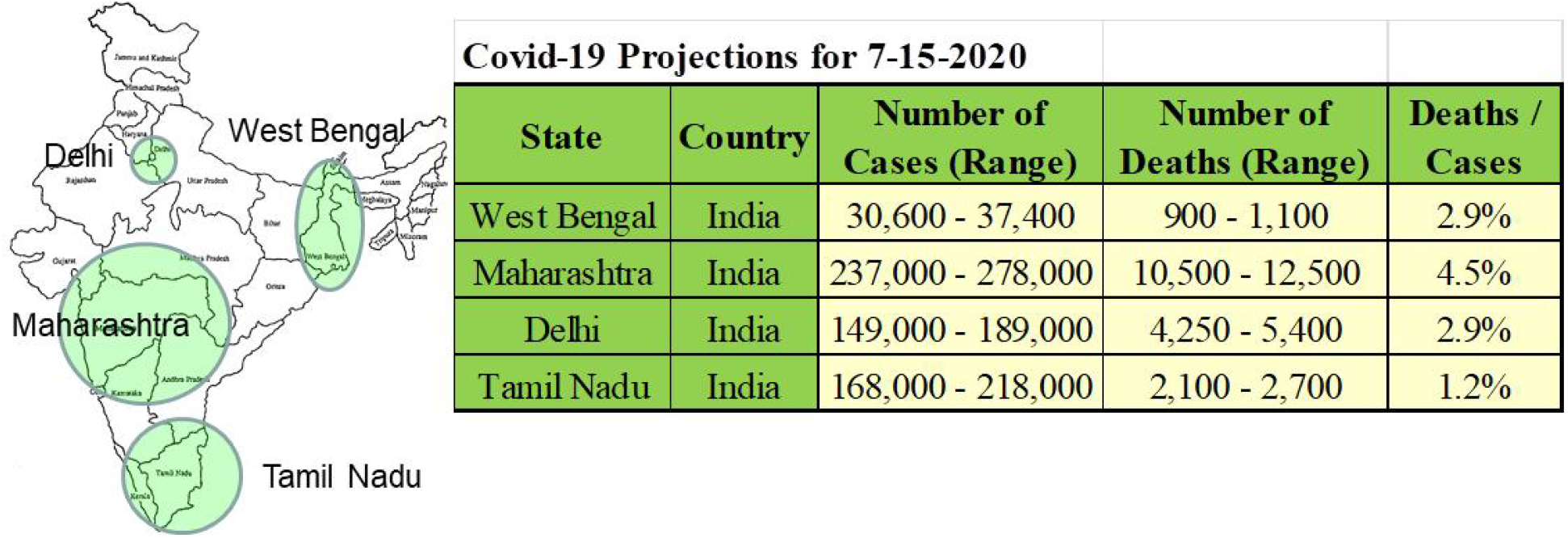
Summary of Covid-19 case and death projections for four states in India

In conclusion, we have used the Basu model to quantitatively examine the Covid-19 case growth in West Bengal, Maharashtra, Delhi and Tamil Nadu in India which are geographically widely separated. In all four states, reopening started before the daily cases have subsided well below the peak due to economic necessity. The consequence is acceleration in Covid-19 case growth which can only be checked by imposing social distancing measures a second time or perhaps multiple times. The case growth during the next thirty days needs to be carefully watched.

The author wishes to acknowledge valuable discussions with P. Soni, L. Gutheinz, P. Hagelstein, W. Grossman, S. Gowrinathan, S. Watanabe, J. Basu, T. Hausken, S. Sheng, T. Mitra, S. Dhawan and M. Genevro.

## Data Availability

The data used is in the paper or referenced in the paper.

https://en.wikipedia.org/wiki/COVID-19_pandemic_in_West_Bengal

https://en.wikipedia.org/wiki/COVID-19_pandemic_in_Maharashtra

https://en.wikipedia.org/wiki/COVID-19_pandemic_in_Delhi

https://en.wikipedia.org/wiki/COVID-19_pandemic_in_Tamil_Nadu

## Notes

### Competing Interest Statement

The authors have declared no competing interest.

### Funding Statement

The work was funded by Sparkle Optics.

### Author Declarations

This mathematical modelling work was approved by Sparkle Optics

## References

1. https://www.cdc.gov/coronavirus/2019-ncov/faq.html#How-COVID-19-Spreads

2. https://en.wikipedia.org/wiki/COVID-19_pandemic_in_New_Zealand

3. Basu, S. (2020, April 28) “A Mathematical Model to Analyze Data on Coronavirus Cases” https://doi.org/10.31224/osf.io/95arg

4. Basu, S. “Modelling to Predict Hospital Bed Requirements for Covid-19 Patients in California”, medRxiv 2020.05.17.20104919; doi: https://doi.org/10.1101/2020.05.17.20104919

5. Basu, S. “Model based comparison of Covid-19 cases in two counties in the silicon valley”, medRxiv 2020.05.26.20114116; doi: https://doi.org/10.1101/2020.05.26.20114116

6. Basu, S. “Model Based Covid-19 Case Studies in the UK, the USA and India”, medRxiv 2020.05.31.20118760; doi: https://doi.org/10.1101/2020.05.31.20118760

7. Basu, S. “Numerical Analysis of Disastrous Effect of Reopening Too Soon in Georgia, USA” medRxiv 2020.07.01.20144667; doi: https://doi.org/10.1101/2020.07.01.20144667

8. https://en.wikipedia.org/wiki/COVID-19_pandemic_in_West_Bengal

9. https://en.wikipedia.org/wiki/COVID-19_pandemic_in_Maharashtra

10. https://en.wikipedia.org/wiki/COVID-19_pandemic_in_Delhi

11. https://en.wikipedia.org/wiki/COVID-19_pandemic_in_Tamil_Nadu

